# Dynamic of humoral response to SARS-CoV-2 anti Nucleocapsid and Spike proteins after CoronaVac vaccination

**DOI:** 10.1101/2021.05.20.21255825

**Authors:** Lucas Bochnia-Bueno, Sergio Monteiro De Almeida, Sonia Mara Raboni, Douglas Adamoski, Ludmilla Louise Moreira Amadeu, Suzana Carstensen, Meri Bordignon Nogueira

**Affiliations:** Virology Laboratory, Federal University of Paraná, General Carneiro, 180, Curitiba, Brazil; Department of Genetics, Federal University of Paraná, Curitiba, PR, Brazil; Post-Graduate Program in Microbiology, Parasitology and Pathology, Federal University of Paraná, Curitiba, PR, Brazil

**Keywords:** SARS-CoV-2, Vaccine, Immunization, Public health, Immunoglobulin G, CoronaVac, Pandemic

## Abstract

**Background:** This study aimed to calculate the seroconversion rate of the CoronaVac vaccine in healthcare workers (HCWs) after immunization.

**Methods:** Serum samples from 133 HCWs from Southern Brazil were collected one day before (Day 0) and +10, +20, +40, + 60, +110 days after administering the vaccine’s first dose. Immunoglobulin G (IgG) was quantified using immunoassays for anti-N-protein (nucleocapsid) antibodies (Abbott, Sligo, Ireland) and for anti-S1 (spike) protein antibodies (Euroimmun, Lübeck, Germany).

**Results:** Seroconversion by day 40 occurred in 129 (97%) HCWs for the S1 protein, and in 69 (51.87%) HCWs for the N protein. An absence of IgG antibodies (by both methodologies), occurred in two (1.5%) HCWs undergoing semiannual rituximab administration, and also in another two (1.5%) HCWs with no apparent reason.

**Conclusion:** This study showed that CoronaVac has a high seroconversion rate when evaluated in an HCW population.

**Funding:** This work was supported by the PROPLAN/Federal University of Paraná, Curitiba-Paraná, Brazil; FINEP, Funder of Studies and Projects, Ministry of Science, Technology and Innovation, Brazil Institutional Network, Project: Laboratories for Diagnostic Tests for COVID-19 (0494/20).

## 1. Introduction

By July 5, 2021, approximately one year after the beginning of the severe acute respiratory syndrome coronavirus 2 (SARS-CoV-2) pandemic, confirmed cases of infection worldwide numbered 183,560,151 people, including 3,978,581 deaths[1]. After the description of this new human coronavirus in December 2019, there was a global effort by researchers, public and private companies in the search for an effective vaccine to control this pandemic[2–4]. These studies resulted in late 2020, with the first doses of immunization in the population, and there are currently 2,988,941,529 doses of the vaccine administered until July 5, 2021[1].

Many SARS-CoV-2 proteins can induce an immune response, amongst them: M (membrane), E (envelope), N (nucleocapsid), and S (spike)[5]. However, the S and N proteins are the most responsive to infection, which induces high titers of anti-SARS-CoV-2 IgM and IgG antibodies. S protein has been more studied for vaccines because it participates in the virus entry mechanism through the connection of the S1 region receptor-binding domain (S1-RBD) in virus particles with the angiotensin-converting enzyme 2 (ACE 2) in the host cell[6,7]. Then, the antibodies binding in this region can cause viral neutralization. Both S and N proteins have also been used for diagnosis, S protein is more specific despite being a more variable portion. In contrast, N protein is a more preserved region, including high homology with N protein SARS-CoV (> 90%), but both may have false-positive results[8]. To evaluate the neutralization antibody activity, the gold-standard assay is the plaque reduction neutralization test (PRNT) that involves the measurement of the ability of patient sera to prevent infection[9]. However, since this assay is time-consuming and requires higher levels of biological safety, multiple groups proposed anti-RBD ELISA assays as a reliable tool to predict neutralization[9–11].

Worldwide efforts resulted in several vaccines against SARS-CoV-2 with distinct antigen platforms systems (non-replicating viral vector, protein subunit, inactivated virus, and mRNA), with the main antigenic focus on S protein[2,3].

The vaccination in Brazil started with CoronaVac (Sinovac Life Sciences, Beijing, China) in January 2021, and until June 2021, two other vaccines come into use in the country. However, CoronaVac (Sinovac Life Sciences, Beijing, China) remains the most administered in Brazilian territory[12], using the inactivated virus as a component of the vaccine[2,3]. In phase I/II studies, this vaccine was safe, tolerable, presented high immunogenicity, and had uncommon adverse reactions. A similar response was observed for both tested concentrations (3 μg and 6 μg), and 97% of seroconversion occurred in the participants with 18-59 ages[11]. In phase III trials, carried out with health care workers, this vaccine presented 50.7%, 83.7%, and 100% efficacy against symptomatic disease, cases requiring assistance, and severe cases, respectively[13]. Phase III also tested some serum samples against the B.1.1.28, gamma (P.1), and zeta (P.2) variants, showing great antibody response[14].

As the vaccine has been administered to people with different ethnicities, comorbidities, and ages, the results of pre-approval clinical trials for its use may not perfectly reflect the response to the vaccine. Thus, vaccine response analyses, either by seroconversion or by neutralizing antibody titration, are essential to assess the possible impacts of this immunization on the population and must be monitored so that the humoral response time can be defined. In this context, this study aimed to identify the seroconversion rate after vaccination with SARS-CoV-2 (CoronaVac) in healthcare workers (HCWs) 40 days after its application.

## 2. Methods

### 2.1 Participants

In total, 170 participants were recruited at the Complexo Hospital de Clínicas, UFPR, Clinical Laboratory, Curitiba, Brazil, during the vaccination of HCWs in this city. The Institutional Ethical Committee approved the study (CAAE: 31687620.2.0000.0096), and all participants signed their consent.

The inclusion criteria were as follows: answering the questionnaire, being vaccinated with two doses of CoronaVac, and providing serum samples. Fourteen participants were excluded because they did not complete the questionnaire. In addition, seven participants took another vaccine, one participant did not have the second dose, and 15 participants did not provide a sample on days 0 (previous vaccination) or +40 (post-vaccination) (Figure 1).

**Figure 1:**
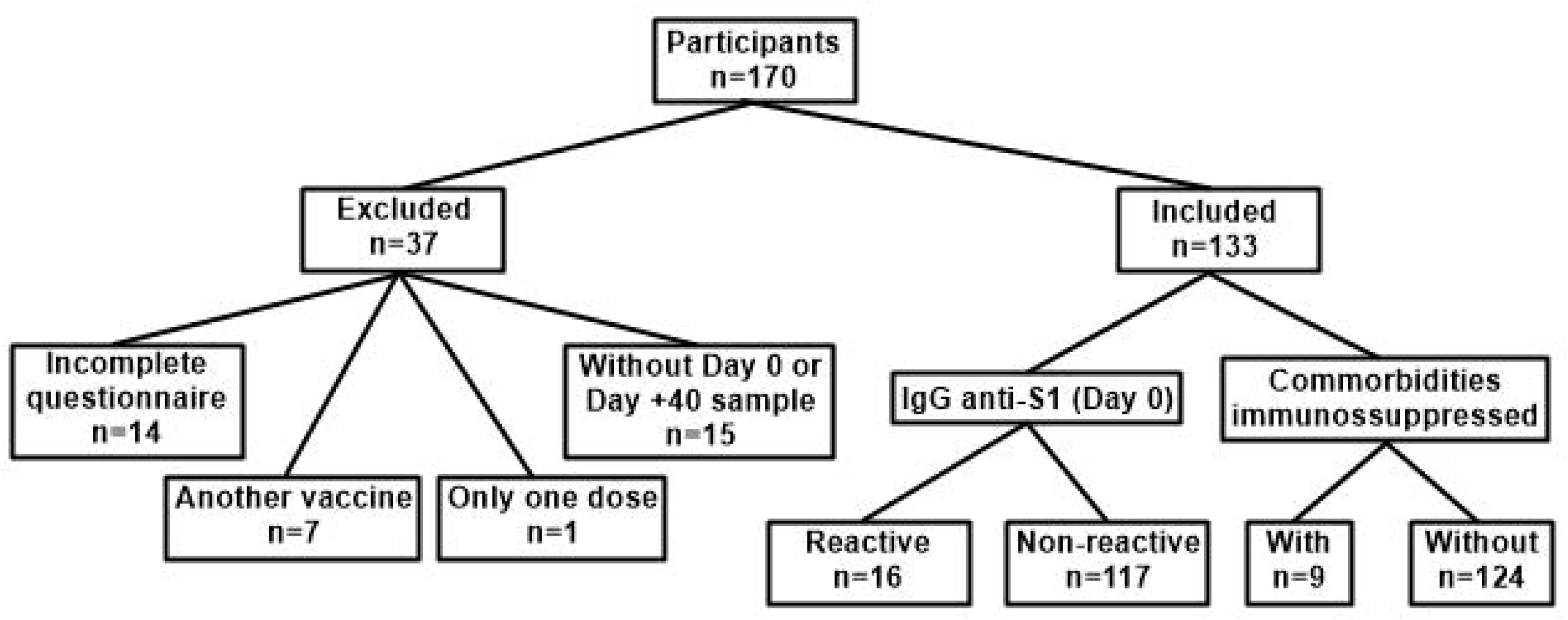
Participants included and excluded in the study and division of groups for analysis. Comorbidities (immunosuppressive) included: Immunosuppressive drugs use, Crohn’s disease, bariatric surgery, HIV and Diabetes.

Serum samples of 133 healthcare workers included in this study were collected on days 0 (previous first dose application), +10, +20, +40, +60, and +110 after the first dose. On day 0 and +40, 133 serum samples were analyzed, and on day +10, +20, +60 and +110, 123, 119, 114 and 132 serum samples were analyzed, respectively. All samples were stored at −20 °C until analysis.

The participants were divided into two groups based on day 0 serology according to anti-spike-1 (anti-S1) immunoglobulin G (IgG)[5,15,16]: *reactive* (n=16) and *non-reactive* (n=117). The participants were also sorted according to the presence of comorbidities into two divisions: *immunosuppressed* (n=9) or *not* (n=124) (Figure 1; Table 1). The immunosuppressed group consisted in participants who presented comorbidity associated with compromised humoral or cellular immune response or those who used immunosuppressive drugs, such as HIV infection, use of chemotherapy or steroids (prednisone at a dose of 20 mg/day or equivalent).

**Table 1:** Demographics characteristics of participants included in the study for each respective group. Information on the handling of special cases: two immunosuppressed (Rituximab 1400 mg/semiannually), one myasthenia gravis (Pyridostigmine 120 mg/day), one Crohn’s disease ostomized twenty-two years ago (Azathioprine 100 mg/day), two participants with bariatric surgery (11 and 12 years), and one HIV+ (Tenofovir 300 mg, Lamivudine 300 mg + Dolutegravir 50 mg/day; CD4^+^ 541/µL). *Comorbidities (immunosuppressive) included: Immunosuppressive drugs use, Crohn’s disease, bariatric surgery, HIV and Diabetes. The patient with Myasthenia gravis is not included here because the treatment used was not immunosuppressive.

|  | IgG Anti-S1 (Day 0) |  | *p* value | Comorbidities immunosuppressive* |  |  |
| --- | --- | --- | --- | --- | --- | --- |
|  | Reactive<br>n (%) | Non-reactive<br>n (%) |  | With<br>n (%) | Without<br>n (%) | *p* value |
| Total | 16 | 117 |  | 9 | 124 |  |
| Female | 13<br>(81.25) | 93<br>(79.49) | 1.0000 | 6<br>(66.67) | 100<br>(80.64) | 0.5636 |
| Median Age<br>(IQR) | 44<br>(25.25 -52.75) | 49<br>(39.50 - 53.50) | 0.2225 | 51<br>(45.50 - 54.50) | 48<br>(38.25 - 53.75) | 0.2297 |

### 2.2 Seroconversion evaluation

Semi-quantitative assays were performed to detect anti-SARS-CoV-2 IgG. For all serum samples, assays used the Chemiluminescent Microparticle Immunoassay (CMIA) Architect-I System for anti-nucleocapsid protein (anti-N) IgG (Abbott, Sligo, Ireland). Additionally, for serum samples from days 0, +40 and +110, assays used the Enzyme-Linked Immunosorbent Assay (ELISA) for IgG anti-S1 spike-protein receptor-binding domain (RBD) (Euroimmun, Lübeck, Germany).

Samples were tested in duplicate, following the manufacturer’s instructions. Results with a variation coefficient greater than 15.0% were repeated.

### 2.3 Statistical analysis

According to the distribution of seroconversion at day +40, the category variables were evaluated using Pearson’s chi-squared test with Yates’ continuity correction. The age variable was evaluated using the Wilcoxon signed rank sum test with continuity correction. Samples paired over time were evaluated using the Friedman ANOVA test (as implemented in the rstatix package), followed by the Wilcoxon signed rank test as a post-hoc pairwise comparison. For samples without multiple observations over time, the Wilcoxon signed rank test was used. All statistical analyses were performed using R (R Core Team). *P* values less than 0.05 were considered significant.

## 3. Results

### 3.1 Seroconversion to S1 protein

Robust production of anti-S1-protein IgG was observed by day +40 in 129 (97%) HCW participants (Figure 2B) by the index test result. Although the reactive (Figure 3D) and non-reactive (Figure 3B) groups had different average index values for S1-protein IgG on day 0 (*p* < 0.0001), on day +40, the average index between the groups was not significantly different (*p* = 0.3704).

**Figure 2:**
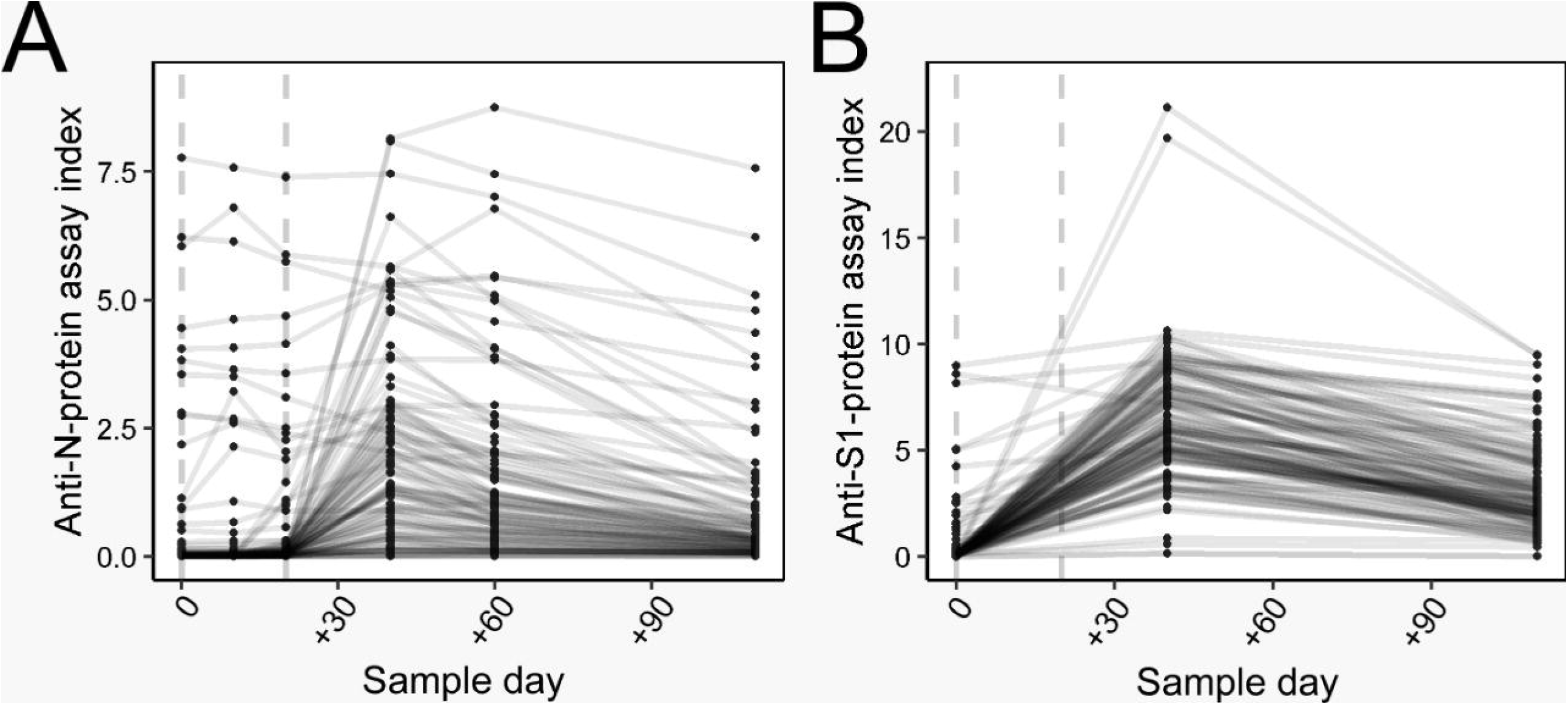
Dynamics of antibody response after vaccine application. The line connecting the dots represents the humoral curve for each participant included in the study. The dotted line represents the days of the vaccine application (two doses). **A** - N-protein IgG evaluation in all participants. **B** - S1-protein IgG evaluation in all participants

**Figure 3:**
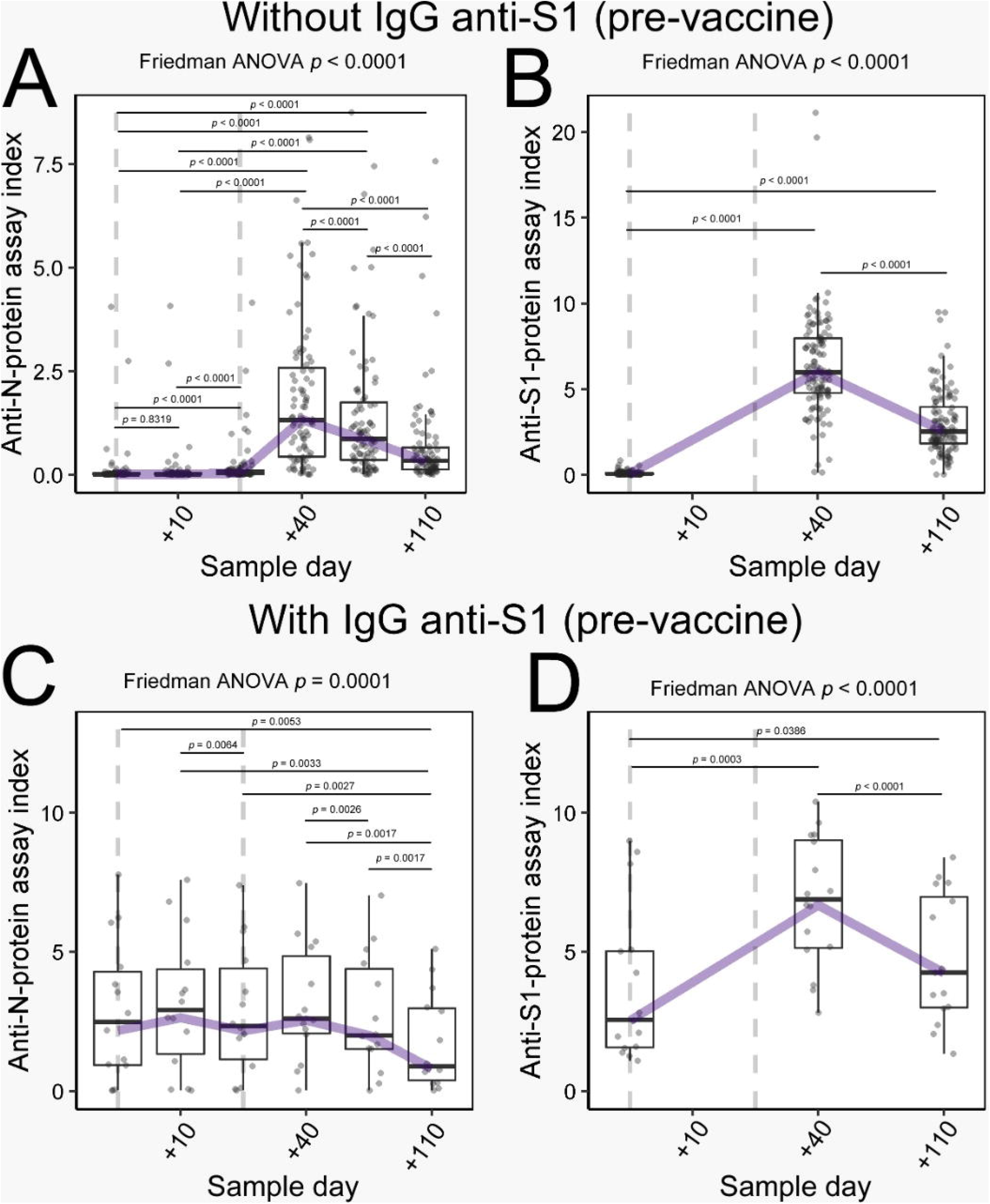
Antibody rates in the S1-protein IgG seroconverted/not seroconverted groups at day 0. Boxplot graph presents median (line dividing the box), interquartile range (box), maximum value (line above the box), and minimum value (line below the box). The line connecting the boxes represents the trend of the data. The dotted line represents the days of the vaccine application (two doses). **A** - N-protein IgG evaluation in S1-antibody nonreactive participants at day 0. **B** - S1-protein IgG evaluation in S1-protein IgG nonreactive participants at day 0. **C** - N-protein IgG evaluation in S1-protein IgG reactive participants at day 0. **D** - IgG anti-S1 protein evaluation in anti-S1 protein IgG reactive participants at day 0.

### 3.2 Seroconversion to N protein

The dynamics of anti-N-protein IgG for all participants are shown in figure 2A. Distributing the data in the division of groups is possible to observe no significant production of the anti-N-protein IgG in non-reactive group participants 10 days after the first vaccine dose (*p* = 0.5027; Figure 3A), and although there was a statistical difference in the sample on day +20 (*p <* 0.0001), there was no apparent seroconversion at that time. By contrast, there was a marked increase in N-protein IgG levels in 69 (51.87%) participants on day +40 (Figure 3A). A significant difference was also observed in the average index for this antibody between the reactive (Figure 3C) and non-reactive groups (Figure 3A): day 0 (*p* < 0.0001) and day +40 (*p* = 0.0657).

### 3.3 Combined response

In the non-reactive group, better-developed antibody responses were observed for N and S1 proteins (*p* < 0.0001; Figure 3A, B), while in the reactive group, the antibody response showed a significant difference (*p* < 0.0001) only for antibodies against S1 protein (Figure 3D), increasing the level of circulating humoral response. No significant changes were observed in IgG anti-N protein analysis for the reactive group at days +10, +20, and +40 (*p* = 0.2231). The antibody index for IgG anti-N and anti-S1 presented at day +40 approximated mean of 2.0 and 6.0, respectively.

Comorbidities were reported by some HCWs, including Crohn’s disease, prior bariatric surgery, HIV+, or diabetes. In general, the participants with comorbidities responded to the vaccine similarly to participants without any comorbidities (Figure 4). However, two cases in the immunosuppressed group did not undergo seroconversion. Furthermore, two other HCWs (not in the immunosuppressed group) did not seroconvert by day +40; both had no apparent cause. These four HCWs without seroconversion were re-evaluated at +60 and +110 days. One participant presented seroconversion of the S1 protein in a sample of +60 days (Figure 5).

**Figure 4:**
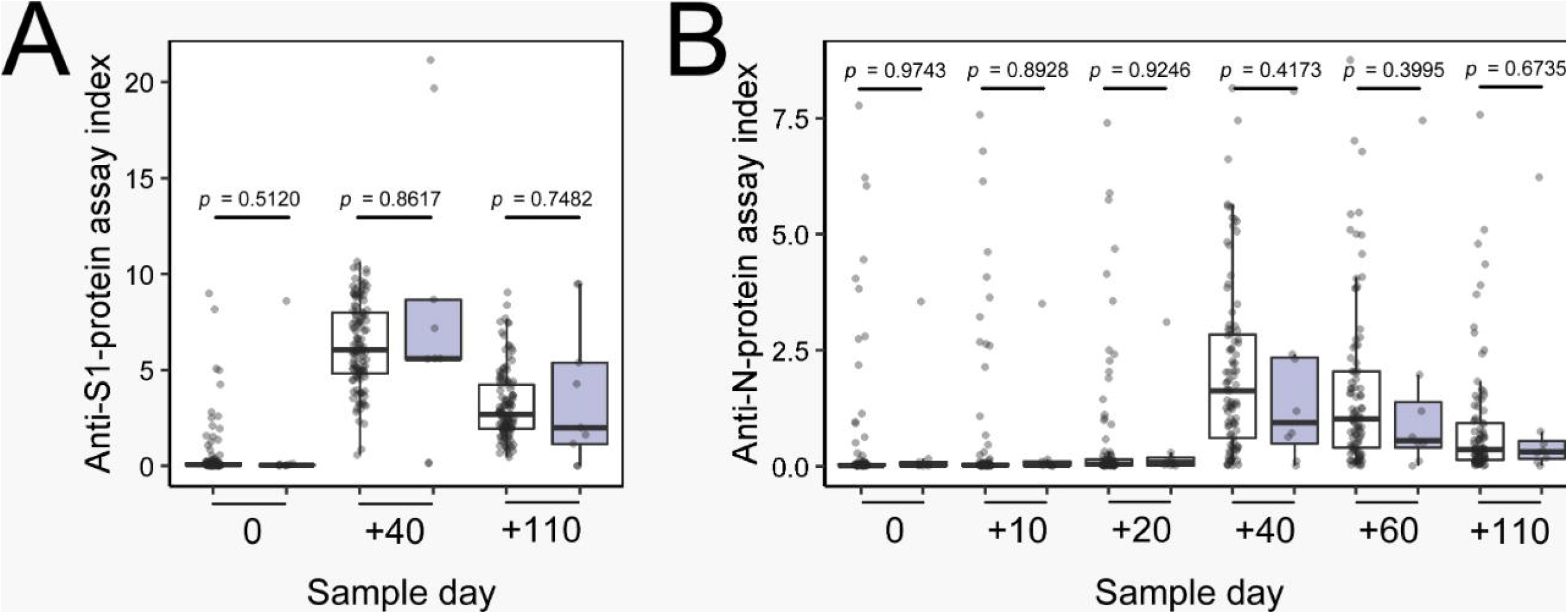
Antibody rates for participants with and without immunosuppression. White boxes indicate non-immunosuppressed participants. Gray boxes indicate immunosuppressed participants. **A** - S1-protein IgG evaluation. **B** - N-protein IgG evaluation.

**Figure 5:**
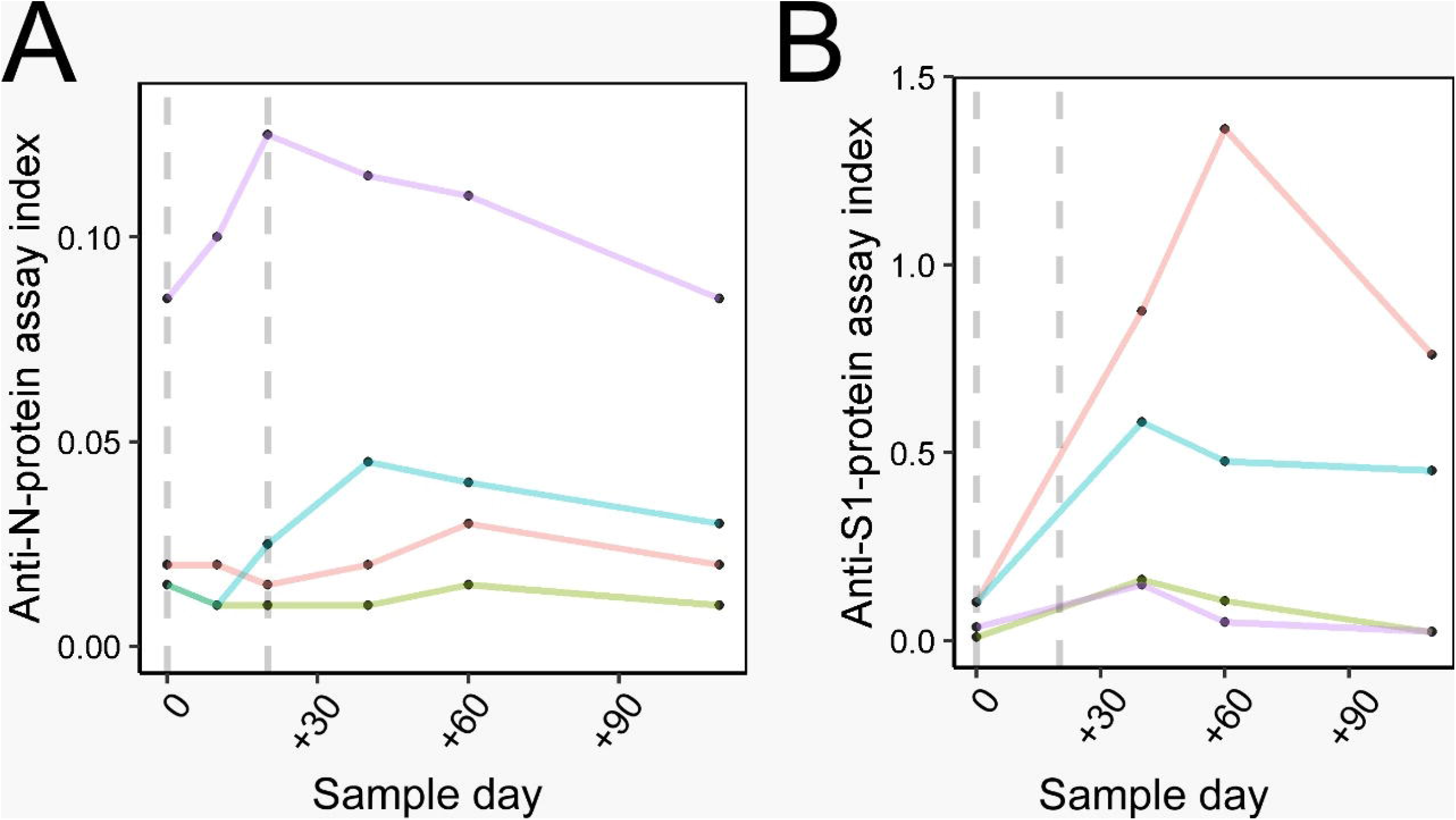
Antibody rates for participants without seroconversion on day +40. Purple and green lines represent the participants with Rituximab treatment. The dotted line represents the days of the vaccine application (two doses). **A** - N-protein IgG evaluation. **B** - S1-protein IgG evaluation.

In the anti-S1 reactive group on day 0, six (37.50%) participants did not have a previous SARS-CoV-2 diagnosis, possibly due to asymptomatic infection. Furthermore, in the anti-S1 non-reactive group, 7 (5.98%) participants had symptoms suggestive of SARS-CoV-2 (fever, dry cough, tiredness, loss of taste or smell, aches and pains, headache, sore throat, nasal congestion, red eyes, diarrhea, or a skin rash)[17], although we did not have information about nasopharyngeal RT-PCR or immunological rapid-test detection. Demographic data according to immunologic response and comorbidities, are shown in Table 1.

### 3.4 Antibodies level maintenance

For both proteins (anti-N and anti-S) in the group without antibodies anti-S at day 0, the antibodies presented index level drop comparing day +40 vs. +110 and index level increase comparing day 0 vs. +110 (all *p*<0.0001) (Figure 3A,B). And in the group with antibodies anti-S at day 0, the antibodies presented index level drop comparing day +40 vs. +110 (N: *p*=0.0017; S: *p*<0.0001), anti-N protein drop comparing day 0 vs. +110 (*p*=0.0053) and anti-S increase comparing day 0 vs. +110 (*p*=0.0386) (Figure 3C,D).

## 4. Discussion

The seroconversion rate of 97% for the anti-S1 IgG observed in HCWs is important data and corroborates the results of phase I/II trials of CoronaVac vaccine[13]. However, it should be noted that the necessary antibody titers for protection are not entirely known. Furthermore, in the clinical trials carried out previously to vaccine registration, the primary outcome was disease severity, so it cannot be affirmed so far whether seroconversion or antibody titers are associated with protection from infection.

Several mutations in the RBD region of the S1 protein have been shown, giving rise to the viral variants of concern, as previously described: gamma (P.1), zeta (P.2), beta (B.1.351), alpha (B.1.1.7), and B.1.325[18–20]. Such mutations confer the potential for the virus to escape the humoral immune response produced due to the disease or to viral vectors or mRNA vaccines[21]. Thus, studies that evaluate vaccine efficacy against these new strains are valuable[22].

Seroconversion rates observed for anti-N protein IgG could be valuable with the emergence of SARS-CoV-2 variants, considering the lower mutation levels in this protein[16], compared to the high mutation levels in the S1 protein[15]. Thus, seroconversion of N-protein antibodies may be an alternative for the vaccine industry to produce efficient vaccines for circulating strains, including those that may arise in the future. However, more studies are needed to understand the impact of antibodies against other viral proteins in the protection against infection.

In this study, there was no difference in the analysis for the anti-N protein IgG in the reactive group, possibly due to the antibody levels present at day 0 in this group; the vaccine has not interfered in the humoral response; the group remained at the same average index. A total of 5.98% of the participants without seroconversion reported they had been previously infected by SARS-CoV 2. All of them presented seroconversion after the complete vaccination. Moreover, whether the person had experienced the disease or not, the levels of antibodies at day +40 post-vaccine were the same. This finding agrees with Krammer et al.[23] in a study of individuals with and without previous COVID-19, given the mRNA vaccine. This same response level implies the same antigen concentration, showing no difference in individual antibody response regardless of the previous infection.

Higher index of anti-S1 antibodies were observed in comparison to the response of anti-N antibodies, corroborating what was exposed by Jiang et al.[8]. The Khoury et al.[24] determination can be used to estimate the level of neutralizing antibodies; for a 50% protection caused by neutralizing antibodies, approximately 20% of the antibody levels observed in the ELISA assays correspond to this level of protection. And for 50% protection in severe cases, only 3% of antibody levels observed in ELISA assays correspond to such protection in severe cases [24]. Therefore, it is possible to estimate the index of neutralizing antibodies in this study.

In participants with immunosuppressive treatment (n=2), the absence of the antibody response was probably due to rituximab having been administered approximately one month before the vaccine. In this situation, as described by Kado et al.[25], there is a significant B lymphocytes decrease. Consequently, there is no production of antibodies until the B lymphocytes recover in 6 to 24 months. In such cases, the response must be evaluated after the repletion time, and re-vaccination considered with medical and clinical endorsement. Two other participants did not seroconvert on day +40. One of these had late-response seroconversion on day +60. No explanation was found for the other case, and more studies are needed to understand what interfered with the immune response.

As with the humoral response developed by other inactivated virus vaccines [26], the dynamics of antibodies produced by CoronaVac in this study shows a peak in the antibody index followed by a sharp drop in that index. It is expected that even with these lower levels, memory B lymphocytes persist for a faster humoral response in cases of reinfection, resulting in less viral activity and minor damage to the host[27]. This lowest observed index has not yet been evaluated to verify whether the remaining humoral response is likely to generate a protective response against an infection.

The immune response developed by vaccination depends not just on antibodies but primarily on neutralizing antibodies[28]. Both natural infection and vaccination act on the immune system in complex ways, stimulating the production of non-neutralizing antibodies (with their specific actions) and TCD4+ and TCD8+ T cells, which also act to protect against COVID-19, as shown by Tarke et al.[28]. That study evaluated the immune response to the SARS-CoV-2 variants and showed that cellular immunity-unlike the humoral response, is little affected by the virus variants. In addition to the specific immune response, innate immunity is another essential protection mechanism against infections[28].

The present study has some limitations: the humoral immunity was studied semi-quantitatively, there was no quantification and titration of anti-SARS-CoV-2 antibodies, and no testing for neutralizing antibodies. The total number of participants was small, and immunosuppressed comorbidities were low in number and had diverse etiologies. More studies are needed to elucidate the vaccine response in these specific groups. However, this is the first study to evaluate the dynamics of IgG anti-N and anti-S1 production after CoronaVac immunization in the community.

The results of seroconversion have shown the importance of two doses for this vaccine as, until the second dose was applied, there was no change in the production of N-protein IgG, as previously described by Zhang et al.[29] in phase I/II tests for this vaccine, with the antibody response detectable just 14 days after the second dose. The second dose is important for several types of vaccines, including mRNA vaccines, as described by Dörschug et al.[30], resulting in a significant increase in antibody levels. Therefore, with SARS-CoV-2, there would be no difference at this point.

In conclusion, significant antibody production was observed 40 days after the first CoronaVac dose in the large majority of study participants, independent of comorbidities. The anti-N protein and anti-S1 protein antibody responses of participants without prior SARS-CoV-2 infection were comparable with those of the previously infected group, in which the immune response was maintained or optimized, with no decrease in levels.

## Data Availability

All data is available with the corresponding author.

## Data Availability

All data is available with the corresponding author.

## Data Availability

All data is available with the corresponding author.

## Acknowledgments

The authors would like to thank all participants who agreed to participate in this study, those involved in the collection and storage of samples, and the Immunochemistry Laboratory section of Complexo Hospital de Clínicas, UFPR, and CAPES.

## Author Contributions Statement

L.B.B.: data collection, data analysis and interpretation, drafting the article, final approval. S.M.A: data analysis and interpretation, drafting the article, critical revision, final approval. S.M.R.: data analysis and interpretation, drafting the article, critical revision, final approval. D.A.: data analysis and interpretation, drafting the article, critical revision, final approval. L.L.M.A.: data collection, drafting the article, final approval. S.C.: data collection, final approval. M.B.N.: conception, data analysis and interpretation, drafting the article, critical revision, final approval, funding acquisition.

